# Efficacy of a nasal spray containing Iota-Carrageenan in the prophylaxis of COVID-19 in hospital personnel dedicated to patients care with COVID-19 disease A pragmatic multicenter, randomized, double-blind, placebo-controlled trial (CARR-COV-02)

**DOI:** 10.1101/2021.04.13.21255409

**Authors:** Juan M. Figueroa, Mónica Lombardo, Ariel Dogliotti, Luis P. Flynn, Robert P. Giugliano, Guido Simonelli, Ricardo Valentini, Agñel Ramos, Pablo Romano, Marcelo Marcote, Alicia Michelini, Alejandro Salvado, Emilio Sykora, Cecilia Kniz, Marcelo Kobelinsky, David Salzberg, Diana Jerusalinsky, Osvaldo Uchitel, CARR-COV2 Group Trial

**Author notes:** Corresponding author: Juan M. Figueroa, Instituto de Ciencia y Tecnología Cesar Milstein (Ciudad Autónoma de Buenos Aires, Argentina). CARR-COV2 Trial Group collaborators are listed at the end of the manuscript.

## Abstract

**Background:** Iota-Carrageenan (I-C) is a sulfate polysaccharide synthesized by red algae, with demonstrated antiviral activity and clinical efficacy as nasal spray in the treatment of common cold. In vitro, I-C inhibits SARS-CoV-2 infection in cell culture.

**Methods:** This is a pragmatic multicenter, randomized, double-blind, placebo-controlled trial assessing the use of a nasal spray containing I-C in the prophylaxis of COVID-19 in hospital personnel dedicated to care of COVID-19 patients.

Clinically healthy physicians, nurses, kinesiologists and others medical providers were assigned in a 1:1 ratio to receive four daily doses of I-C spray or placebo for 21 days.

The primary end point was clinical COVID-19, as confirmed by reverse-transcriptase–polymerase-chain-reaction testing, over a period of 21 days. The trial is registered at ClinicalTrials.gov (NCT04521322).

**Findings:** A total of 394 individuals were randomly assigned to receive I-C or placebo. Both treatment groups had similar baseline characteristics.

The incidence of COVID19 was significantly lower in the I-C group compared to placebo (1·0% vs 5·0%) (Odds Ratio 0·19 (95% confidence interval 0·05 to 0·77; p= 0·03). Workday loss in placebo group compared to I-Cc were 1.6% days / person (95% ci, 1.0 to 2.2); p <0.0001

There were no differences in the incidence of adverse events across the two groups (17·3% in the I-C group and 15·2% in the placebo group, p= 0·5).

**Interpretation:** I-C showed significant efficacy in preventing SARS-Cov-2 infection in hospital personnel dedicated to care patients with COVID-19 disease.

**Research in context:** *Evidence before this study:* We searched PubMed for research articles published up to February 14, 2021, with no language restrictions, using the terms “SARS-CoV-2” or “COVID-19”, “prevention”, “clinical trial”, and “prophylaxis”. Except for studies on vaccines we only found three peer-reviewed publications available on the efficacy of Hydroxycholoquine to prevent COVID-19 disease in individuals at risk of exposure. Hydroxychloroquine did not prevent COVID-19 used as pre or postexposure prophylaxis. We also did not find results from clinical trials on the efficacy of carrageenan in the prevention or treatment of cOVID-19.

*Added value of this study:* We report the clinical efficacy of a nasal spray with Iota-Carrageenan for the prevention of COVID-19 disease in a randomised, double-blind, placebo-controlled, multicentre study in República Argentina, including 394 participants.

*Implications of all the available evidence:* A simple intervention such as the administration of a nasal spray with Iota-Carrageenan, in addition to hand hygiene, use of personal protective equipment and social distancing, could provide additional protection until vaccines can be administered to the majority of the population.

## Introduction

A novel coronavirus, severe acute respiratory syndrome coronavirus 2 (SARS-CoV-2), was first identified in December 2019 as the cause of a respiratory illness designated Coronavirus disease 2019, or COVID-19. Current available evidence shows that COVID-19 virus is transmitted between people through close contact and droplets. Being in close contact with infected individuals is therefore a risk factor to contract COVID19. Unvaccinated health care providers, who are in close contact with COVID-19 patient are therefore at an increased risk for COVID19. This inevitably places unvaccinated health and other hospital workers at a high risk of infection. Recent COVID19 vaccine developments have shown a high efficacy at preventing COVID19 (1,2), and vaccination rate among healthcare workers in high income countries has grown steadily over the first quarter of 2020 (3-6). Nevertheless, vaccine production challenges, distribution delays and global vaccine access have once again highlighted global inequality. The need to develop additional low cost interventions to mitigate the risk of contracting COVID19 among unvaccinated healthcare providers in particularly important for the global South, whore vaccination rate among healthcare providers remains low. The existence of a prophylactic intervention against this disease (except for vaccines already available) remains unknown.

Iota-carrageenan -a sulfated polysaccharide found in some species of red seaweed (Chondrus crispus)-has demonstrated antiviral activity against respiratory and other viruses in cell culture and in animal models.(7-10) Iota-carrageenan inhibits viruses based on its interaction with the surface of viral particles, thus preventing them from entering cells and trapping the viral particles released from the infected cells. *In vitro* and *in vivo* studies have demonstrated the effectiveness of Iota-Carrageenan against several respiratory viruses such as HRV, influenza A and common cold Coronavirus. Carrageenan is generally recognized as safe for use in food and topical applications.

Because the primary site of infection and replication of most cold-causing viruses is the nasal mucosa, it has been hypothesized that early and targeted treatment of the nasal mucosa with Iota-Carrageenan may block viral entry on that level and interfere locally with the propagation of viral replication.

In three randomized clinical trials (two in adults and one in children) that compared Iota-Carrageenan nasal spray with saline solution (placebo) there were strong indications of efficacy, including significantly reduced cold symptoms;(11) positive effects on symptoms in patients in whom less co-medication or no co-medication was used;(12) significantly reduced viral loads;(11-13) and faster reduction of common cold symptoms.(12-13) Treatments were safe and well tolerated.(11-13) In cell culture Iota-Carrageenan has demonstrated antiviral activity against SARS-CoV-2 virus (14,15) and SARSCoV-2 Spike Pseudotyped Lentivirus (SSPL).(16) Taking into account that the concentrations found to be active *in vitro* against SARS-CoV-2 may be easily achieved by the application of nasal sprays already marketed in several countries,(14) and that during the first days of disease the virus is localized mainly in the nasal cavity and the nasopharynx,(17) we hypothesized that a nasal spray with Iota-Carrageenan could potentially be used as preexposure and during exposure prophylaxis, to prevent symptomatic infection in health workers at high risk of COVID19 infection.

## Methods

### Study design and participants

We conducted a pragmatic randomized, placebo-controlled clinical trial to determine if a nasal spray with Iota-Carrageenan can prevent COVID-19 infection in healthcare workers caring for COVID-19 patients. This study was carried out when vaccination plans had not yet begun in Argentina. We randomly assigned participants in a 1:1 ratio to receive either Iota-Carrageenan or placebo. Trial enrollment began on July 24, 2020. Health and other hospital workers attending patients with a positive polymerase-chain-reaction (PCR) assay for SARS-CoV-2 admitted in hospitals were eligible. This trial was approved by the institutional review board and by the ethics committees of each participating center, and participants provided written consent prior to participation.

We included physicians, nurses, kinesiologists and other hospital workers with no history of clinical SARS-CoV-2 infection, who performed medical care in a COVID19 hot zone in the hospital, and were therefore exposed daily to patients with COVID-19. Participants were excluded if (a) they were younger than 18 years of age, (b) participated in any other clinical trial of an experimental treatment for COVID-19, (c) had not entered an area with new patients admitted for COVID-19 in the last 24 hours, (d) did not have a cell phone for remote monitoring, (e) reported hypersensitivity or known allergy to any component of the product, or (f) were pregnant or lactating. Additionally, medical personnel under suspicion of COVID-19, COVID-19 history or with COVID-19 antibodies found in a previous routine screening were deemed uneligible to participate in this study.

### Randomisation and masking

Randomisation occurred at the coordinator center. It was generated a permuted-block randomization sequence using sized blocks of 8. A research pharmacist sequentially assigned participants to either of the groups. The assignments were concealed from investigators and participants (double blind).

### Procedures

Participants were instructed to self-administer 1 puff (0·10 mL) of trial medication to each nostril 4 times per day. Trial medication was either Iota-Carrageenan nasal spray (1·70 g Iota-Carrageenan/L in 0·9 % NaCl) or placebo (0·9 % NaCl). The inhaler bottles containing the active intervention or placebo were identical and odorless. The active drug is approved for use in this dosage by regulatory authorities and available on the market. Both the active sprays and the placebos were provided free of charge by the manufacturer.

Treatment was to be mandatory for 21 days. Participants continued to adhere to handwashing, use of personal protection equipment, physical distancing and general guidelines in compliance with regulations from health authorities. Follow-up was measured on 21th day.

#### Outcomes

The primary outcome was prespecified as symptomatic illness confirmed by a positive molecular assay (PCR) at a local testing facility (using a protocol-defined acceptable test). COVID-19 –related symptoms were the self-reported (any of them) presence of cough, shortness of breath, or difficulty breathing, fever, chills, rigors, myalgia, headache, sore throat, new olfactory and taste disorders, diarrhea and/or vomiting.

In any case, the participants received a daily message on their phone, with a structured questionnaire with the symptoms that should be reported. These symptoms were reported to the center’s main investigating doctor, who confirmed the clinical suspicion and requested the test to determine the presence of COVID-19 disease.

#### Statistical analysis

We estimated that 200 participants would need to be enrolled in each group to give the trial approximately 80% power, at two-sided type I error rate of 5%, to show that COVID-19 would be 50% lower in active treatment group than in the placebo. The strength of association was expressed as a relative risk reduction and its 95% confidence intervals (95% CI). Proportions were compared with the chi-square test or Fisher’s exact test, and the continuous quantitative variables with the Student’s t test. We conducted all analyses with SAS software, version 9·4 (SAS Institute), according to the intention-to-treat principle, with two-sided type I error with an alpha of 0·05.

## Results

From July 24, 2020, to December 20, 2020, a total of 400 hospital workers were enrolled and underwent randomization at 10 hospitals in Argentina.

Six participants were excluded from the final analysis because they had symptoms suggestive of COVID-19 at the time of randomization. Of the remaining 394 participants, 196 had been assigned to receive Iota-Carrageenan and 198 to receive placebo.

Thirteen individuals in Iota-Carrageenan group and 14 in placebo group withdrew consent before day 21 and did not provide information about their health status. The mean age of participants was 38.5±9 years old, and 75.1% were female gender (Table 1).

**Table 1:** Baseline characteristics of the intention-to-treat population

|  | I-C<br>(n= 196) | Pacebo<br>(n= 198) | *p* value |
| --- | --- | --- | --- |
| Sex |  |  |  |
| Female | 141 (71.9%) | 155 (78.3%) | 0.1 |
| Male | 55 (28.1%) | 43 (21.7%) |  |
| Age, years | 38.3 (10.1) | 38.8 (9.2) | 0.8 |
| Ethnic origin |  |  |  |
| White and latino | 196 (100%) | 198 (100%) | 1.0 |
| Physicians | 97 (49.5%) | 95 (48.0%) | 0.8 |
| Nurses | 50 (25.5%) | 62 (31.3%) | 0.2 |
| Technicians | 24 (12.2%) | 26 (13.1%) | 0.8 |
| Others medical providers | 22 (11.2%) | 18 (9.1%) | 0.5 |
| Co-morbidities |  |  |  |
| No co-morbidities | 157 (80.1%) | 147 (74.2%) | 0.2 |
| Chronic pulmonary disease | 6 (3.0%) | 7 (3.5%) | 1.0* |
| High blood pressure | 9 (4.6%) | 10 (5.1%) | 0.8 |
| Obesity | 7 (3.6%) | 13 (6.6%) | 0.2 |
| Severe obesity | 0 (0%) | 0 (0%) |  |
| Hypothyroidism | 12 (6.1%) | 9 (4.5%) | 0.5 |
| Smoking | 1 (0.5%) | 3 (1.5%) | 0.6* |
| Type 2 diabetes mellitus | 0 (0%) | 1 (0.5%) | 1.0* |
| Cancer | 0 (0%) | 0 (0%) |  |
| Chronic kidney disease | 0 (0%) | 0 (0%) |  |
| Down syndrome | 0 (0%) | 0 (0%) |  |
| Heart disease | 0 (0%) | 0 (0%) |  |
| Immunocompromised state | 0 (0%) | 0 (0%) |  |

Data are n (%), mean (SD)
I-C: Iota carrageenan. Obesity (body mass index of 30 Kg/m<sup>2</sup> or higher but <40 kg/m<sup>2</sup>).
\* Fisher's exact test

Forty three participants underwent a PCR test for presenting symptoms compatible with COVID-19 (Table 2), 31 were negative (7·6% in the Iota-Carrageenan group and 8.6% in the placebo group; *p*= 0·8).

**Table 2:**
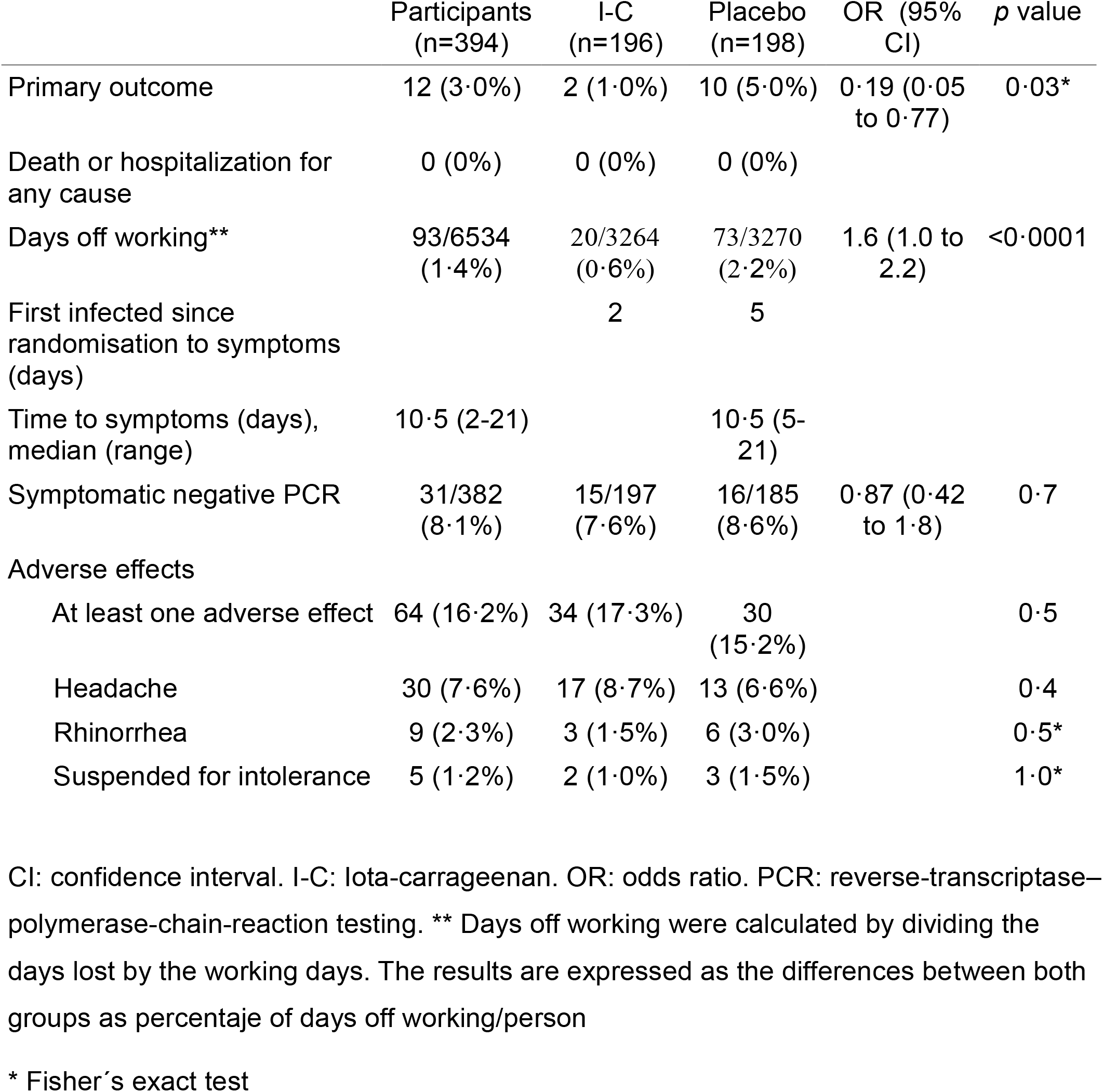
Findings

|  | Participants<br>(n=394) | I-C<br>(n=196) | Placebo<br>(n=198) | OR (95%<br>CI) | p value |
| --- | --- | --- | --- | --- | --- |
| Primary outcome | 12 (3.0%) | 2 (1.0%) | 10 (5.0%) | 0.19 (0.05<br>to 0.77) | 0.03* |
| Death or hospitalization for<br>any cause | 0 (0%) | 0 (0%) | 0 (0%) |  |  |
| Days off working** | 93/6534<br>(1.4%) | 20/3264<br>(0.6%) | 73/3270<br>(2.2%) | 1.6 (1.0 to<br>2.2) | <0.0001 |
| First infected since<br>randomisation to symptoms<br>(days) |  | 2 | 5 |  |  |
| Time to symptoms (days),<br>median (range) | 10.5 (2-21) |  | 10.5 (5-21) |  |  |
| Symptomatic negative PCR | 31/382<br>(8.1%) | 15/197<br>(7.6%) | 16/185<br>(8.6%) | 0.87 (0.42<br>to 1.8) | 0.7 |
| Adverse effects |  |  |  |  |  |
| At least one adverse effect | 64 (16.2%) | 34 (17.3%) | 30<br>(15.2%) |  | 0.5 |
| Headache | 30 (7.6%) | 17 (8.7%) | 13 (6.6%) |  | 0.4 |
| Rhinorrhea | 9 (2.3%) | 3 (1.5%) | 6 (3.0%) |  | 0.5* |
| Suspended for intolerance | 5 (1.2%) | 2 (1.0%) | 3 (1.5%) |  | 1.0* |

CI: confidence interval. I-C: Iota-carrageenan. OR: odds ratio. PCR: reverse-transcriptase–polymerase-chain-reaction testing. \*\* Days off working were calculated by dividing the days lost by the working days. The results are expressed as the differences between both groups as percentage of days off working/person
\* Fisher's exact test

Overall, new COVID-19 (symptomatic with PCR-confirmed) developed in 12 of 394 participants (3.04%) during the 21 days of follow-up (Table 2).

The incidence of COVID-19 differs significantly between those receiving the nasal spray with Iota-Carrageenan (2 of 196 [1·0 %]) and those receiving placebo (10 of 198 [5·0 %]) (Odds Ratio 0.19 (95% confidence interval 0.05 to 0.77; p=0.03). Business day losses censored at day 21 were lower in I-C group (0.5% and 2.0%; p< 0.0001). In sensitivity analysis in which we removed from our analyses individuals who presented symptoms before 7 days after randomization, the risk reduction was 95% (95% CI, 6.0% to 99.7%), *p*= 0.04. OR: 0.05 (95% CI, 0.003 to 0.9), p=0.04.

Days off work in placebo group compared to I-C were 1.6% days / person (95% CI, 1.0 to 2.2); p <0.0001

In the Iota-Carrageenan group and placebo group, 17.3% and 15.2%, respectively, reported at least one adverse effect (p = 0.5)

## Discussion

The results of this study show that the Iota-Carrageenan nasal spray is safe and effective to prevent COVID-19 disease in hospital workers providing care for COVID-19 patients. In our study we identified a risk reduction greater than 80%. This finding is particularly relevant as until now the only prophylactic interventions with demonstrated efficacy are vaccines who are not yet accessible worldwide. In facts, vaccination rates among healthcare workers remain particularly low, specially in the global south.

There has been growing interest in the potential efficacy of drugs with demonstrated in vitro efficacy. During the early days of the COVID-19 pandemic, there was an increased attention to the use of hydroxychloroquine, an agent that was active in vitro but did not prevent COVID-19 when used as pre or postexposure prophylaxis(18-20). With at least two registered clinical trials as of February 2021 (Argentina and Austria), Iota-Carrageenan is being proposed as a potential efficacious prophylactic drug. The nasal spray with Iota-Carrageenan has already shown clinical efficacy in diseases of the upper airways produced by viruses against which Iota-Carrageenan had demonstrated efficacy in vitro. Additionally, Iota-Carrageenan’s in vitro efficacy was shown *in vitro* concentrations equal to and up to 100 times lower than those estimated to be reached in the nasal cavity with the use of sprays available in different countries with standard dosages. We have recently repeated the in vitro study of the effect of carrageenan spray on SARS-COV-2 infection in cultures of a human respiratory epithelium cell line (Calu-3) observing the same inhibitory effect as in Vero cells (submitted).

Our study have some limitations. First, we included apparently healthy individuals without confirmation by PCR test. Second, those who remained asymptomatic were also not tested. Third, we do not know the exposure dose of each participant, although, the number of active principle and placebos administered in each participating center were identical. The devastating urgency of the COVID-19 pandemic requires a simple and pragmatic design trial with the ability to give, in this context, a quick and efficient answer. This is particularly important considering that health providers are overworked and extremely busy, and a higher burden associated with completing numerous data would have resulted in low study compliance.

Our study has also a number of strengths. First, we chose healthcare and other hospital workers to participate in this research as a simple and easy-to-follow model. Second, enrollment took place during a high rate of community transmission in Argentina, therefore, our participants were also exposed outside the hospital. Third, and following the pragmatic nature of this randomized controlled clinical trial and according to the regulations established by the Nation Ministry of Health, we performed only one PCR test between 48 and 72 hours after the onset of symptoms, assuming that a negative first test may not have been enough, nor we have carried out antibody dosages to confirm the disease. Finally, a small number of individuals were lost to follow up (6.8%). In sensitivity analysis where it was hypothesized that the 13 lost individuals from the Iota-Carrageenan group were infected, and that the 14 lost individuals from the placebo group were not infected, no differences were found in infection rates of both groups (*p*= 0.3).

## Conclusions

The nasal spray with I-C showed significant efficacy in preventing SARS-Cov-2 infection in personnel dedicated to care patients with Covid-19 disease. This finding should be replicated in future clinical trials.

## Data Availability

The data supporting the study findings are available within the paper.

## Acknowledgements

We thank the participating hospitals, their staff and persons included in this study.

## Author contributions

JMF, ML, AD, LPF were responsible for the study concept, design, acquisition of data, verification of the underlying data and drafting the manuscript. ML and AD performed statistical analyses. RPG and GS participated in data interpretation, writing the manuscript and agreed on the decision to publish. All authors reviewed, critically revised and approved the final version of the manuscript.

## Conflict of Interest

JMF and ML reports personal fees from Laboratorio Pablo Cassará, outside the submitted work.

Clinical trials registration NCT04521322

---

CARR-COV2 Trial Group: M.Jimena Ortega^2^, Cristina Soler Riera^2^, Ana Cajelli^8^, Fernando Ross^8^, Mirta Gutiérrez^8^; Viviana Jalife^9^, Mariel Trinidad^9^, Paula Bellagamba^9^; Teresa Corallo^10^, Daniel Lamberti^10^; Pablo Oyhamburu^11^, Yael Gonzalez^11^, Carmen Rios^11^, Glenda Ernst^11^, Victor Ikeda^12^, Carolina Osuna^12^, Juan Sang^13^, Natalia Judis^13^, Sofía Golé^15^, Lorena Itati Ibanez^18^, Dr. Ricardo Reisin^11^

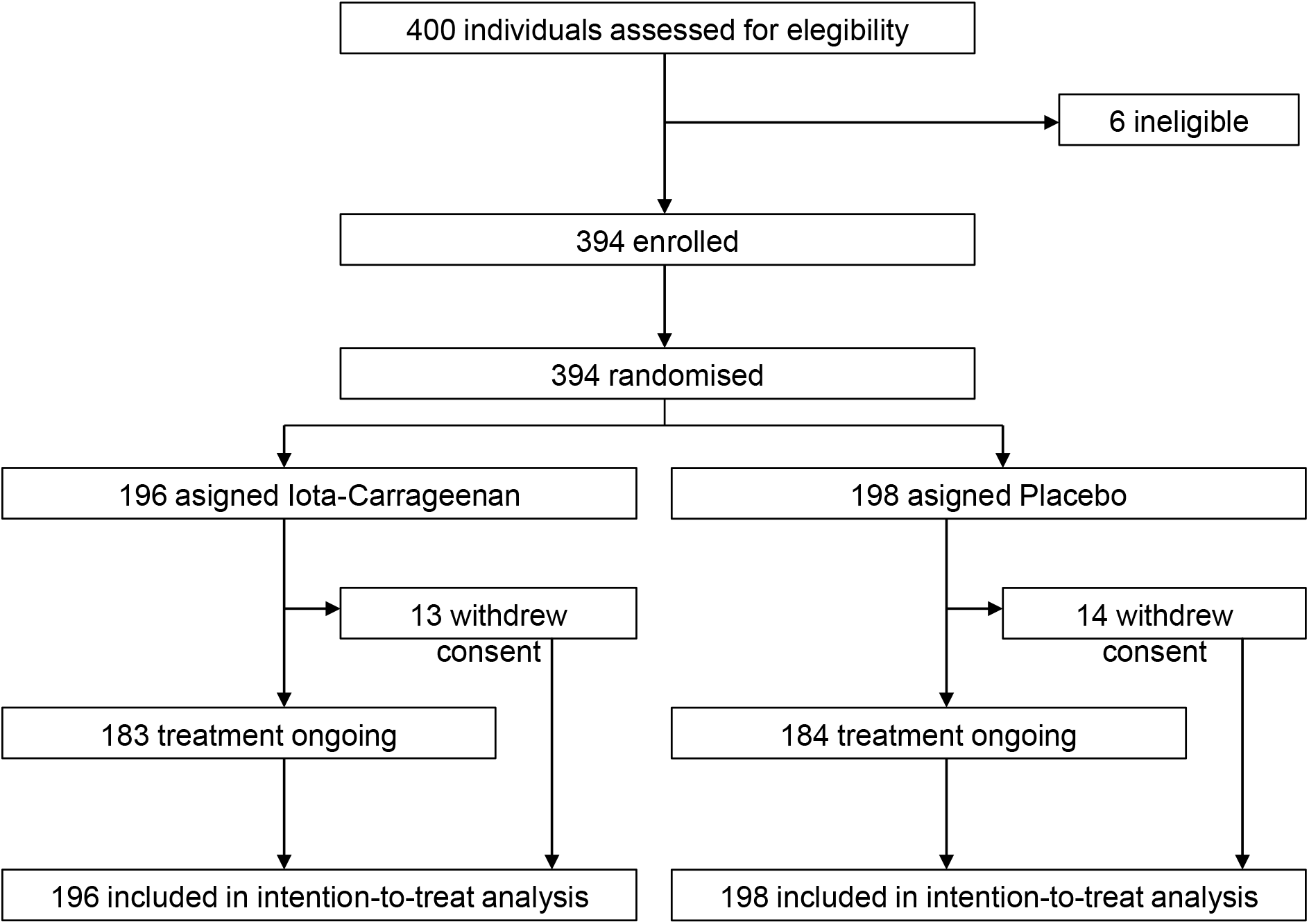

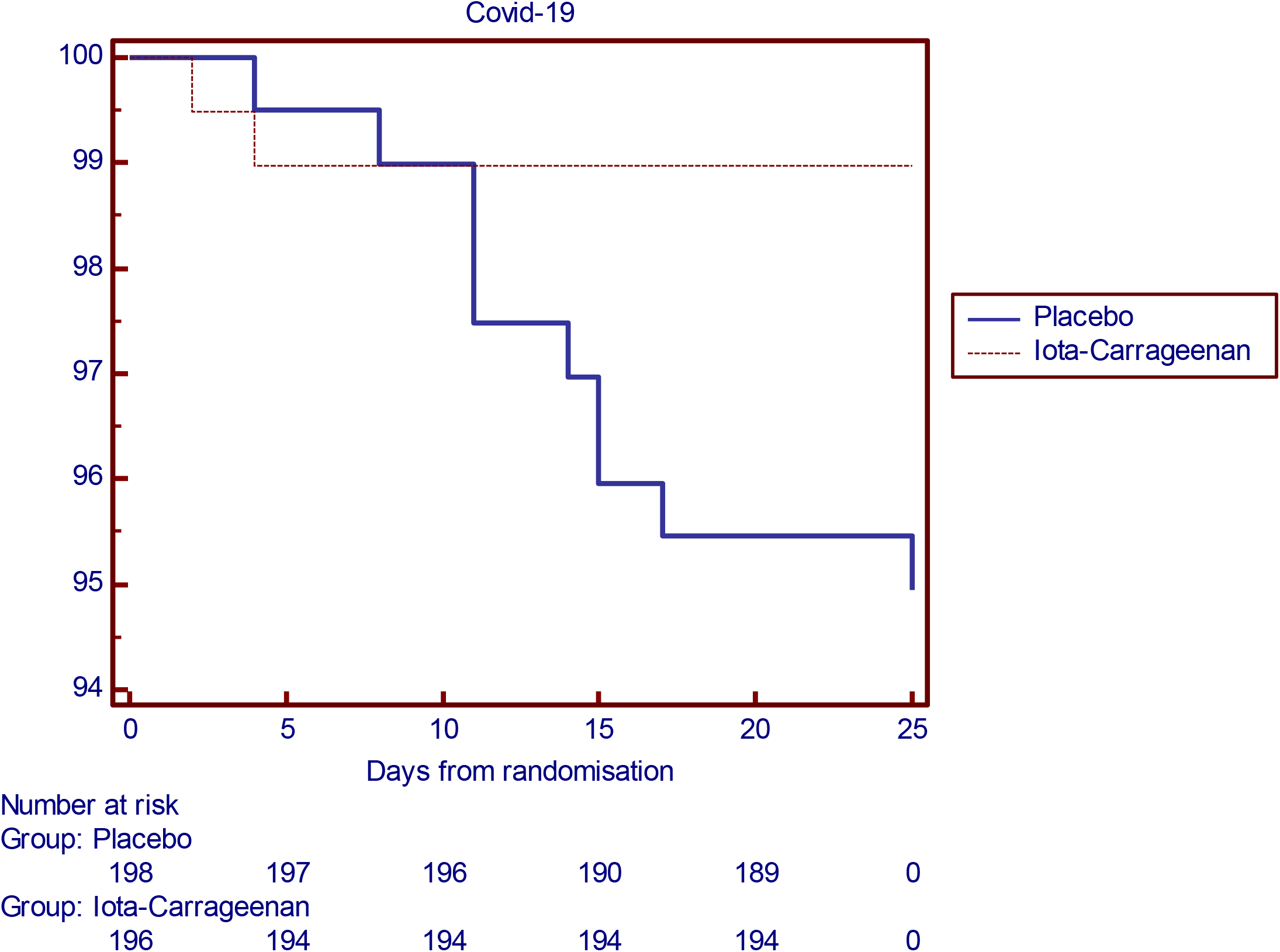

